# Polygenic coronary artery disease association with brain atrophy in the cognitively impaired

**DOI:** 10.1101/2022.02.11.22270852

**Authors:** Eric de Silva, Carole H Sudre, Josephine Barnes, Marzia A Scelsi, André Altmann, the Alzheimer’s Disease Neuroimaging Initiative

## Abstract

The role cardiac function plays in the predilection for, and progression of, Alzheimer’s disease (AD), is complex and unclear. While a number of low-frequency genetic variants of large effect-size have been shown to underlie both cardiovascular disease and dementia, recent studies have highlighted the importance of common genetic variants of small-effect size, which, in aggregate, are embodied by a polygenic risk score (PRS). In this study we aim to investigate the effect of polygenic risk for coronary artery disease (CAD) on brain atrophy in AD using whole brain volume (WBV) and put our findings in context with the polygenic risk for AD and presumed small vessel disease as quantified by white matter hyperintensities (WMH). We used 730 subjects from the ADNI database to investigate PRS effects (beyond APOE) on whole brain volumes, total and regional WMH and amyloid beta across diagnostic groups. In a subset of these subjects (N=602) we utilise longitudinal changes in whole brain volume over a maximum of 24 months using the boundary shift integral approach. Linear regression and linear mixed effects models were used to investigate the effect of WMH at baseline as well as AD-PRS and CAD-PRS on whole brain atrophy and whole brain atrophy acceleration, respectively. All genetic associations were examined under oligogenic (p=1e-5) and the more variant-inclusive polygenic (p=0.5) scenarios. Our results suggest no evidence for a link between PRS score and markers of AD pathology at baseline (when stratified by diagnostic group). However, both AD-PRS and CAD-PRS were associated with longitudinal decline in WBV (AD PRS t=3.3, P_FDR_=0.007 over 24 months in healthy controls) and surprisingly, under certain conditions WBV atrophy is statistically more correlated with cardiac PRS than AD PRS (CAD PRS t=2.1, P_FDR_=0.04 over 24 months in the MCI group). Further, in our regional analysis of WMH, AD PRS beyond APOE is predictive of white matter volume in the occipital lobe in AD subjects in the polygenic regime. Finally, the *rate of change* of brain volume (or *atrophy acceleration*) may be sensitive to AD polygenic risk beyond APOE in healthy individuals (t=2, p=0.04). For subjects with mild cognitive impairment (MCI), beyond APOE, a more inclusive polygenic risk score including more variants, shows CAD PRS to be more predictive of WBV atrophy, than an oligogenic approach including fewer larger effect size variants.

## 1. Introduction

It is estimated that more than one million people in the UK, and over 44 million individuals globally^1^ are living with dementia. Alzheimer’s disease (AD) is the most common form of dementia and is usually diagnosed in the elderly (over the age of 65 years). There are many symptoms associated with AD and these include changes in memory, language and personality^2^.

Beyond cognitive testing there has been a large focus on the use of brain imaging to help diagnose and track the disease. In particular, a decrease in brain volume and the build-up of protein in the form of amyloid plaques (between neurons) and misfolded neurofibrillary tangles of hyperphosphorylated tau (within neurons) are seen in brain imaging studies and observed in post-mortem examinations of AD patients^3^. It is thought that the amyloid and tau aggregations contribute to the death of neuronal cells resulting in a reduction in regional grey-matter volume and neuronal connectivity^4^. Surprisingly, years of clinical trials involving pharmacological interventions targeting these protein deposits have largely been unsuccessful^5^, possibly because they are formed as a result of earlier pathological changes. Another explanation may be that drug interventions, thus far, have all been administered too late in the disease life-course to be effective, and that intervention earlier in life may be required. Current symptomatic treatments of AD symptoms come from acetylcholinesterase inhibitors and glutamate blockers which serve, in some cases, to reduce disease severity, but are by no means curative^6^. At the time of writing the monoclonal antibody aducanumab, which targets beta-amyloid plaques, had just been approved for clinical use in the United States (see www.fda.gov).

AD is often diagnosed alongside vascular dementia in what is called mixed dementia^7^. Vascular dementia (itself the second most common form of dementia following AD) is the result of reduced blood flow to brain cells resulting in cell-death. This can be caused by cerebral small vessel disease, resulting in subcortical vascular dementia, which affects vessels deep in the brain. One imaging marker of small vessel disease is MRI-visible white matter changes, or, white matter hyperintensities (WMHs). WMHs are known to increase with age^8,9^, but are also strongly associated with AD^10^. WMHs have also been associated with a range of other pathologies, including amyloid angiopathy, arteriosclerosis, axonal loss, blood–brain barrier leakage, demyelination, gliosis, hypoperfusion, hypoxia and inflammation^11^.

In coronary artery disease (CAD) plaques aggregate in blood vessels that feed the heart oxygen and nutrients. In the extreme, this can lead to angina and heart attack, but a smaller prolonged reduction in cardiac function may be responsible for cerebral hypoperfusion. CAD has a strong genetic basis, being ∼50% heritable with ∼60 genetic loci identified^12^. However, the relationship between the genetic contribution to heart health and AD remains largely unexplored.

While age is the most salient factor in AD risk, there is, alongside environmental and lifestyle factors, a genetic component underlying both AD and CAD. In AD, a small number of cases (<5%) are due to autosomal dominant early-onset AD for which there are a number of rare, large effect-size genetic variants that contribute to the pathology^13^. These include mutations that result in abnormal protein products of amyloid precursor protein (APP), or in the genes that code for the enzymes that alter the breakdown of APP, both of which may result in an increase in amyloid plaques.

Most AD cases are sporadic and late-onset (typically found in those aged 65 and older) where heritability is estimated to be between 60% and 80%^14^. Here, a number of identified common variants, most notably the e4 allele of the *APOE* gene which accounts for ∼5% of AD heritability, plus about 20 additional loci, account for ∼30% AD heritability^15^. It is likely that the remaining heritability is the result of the combination of a great many (1000s to 100,000s) of common variants, each contributing a very small effect.

As a result of these findings, much research has been performed to establish how to best capture a composite measure of these many common variants, that individually have such a small effect. Polygenic Risk Scores (PRSs) offer a way of doing this and have become increasingly used following the many large Genome Wide Association Studies (GWAS) which show associations between common genetic variants and diseases. The PRS sums up the effect size^16^ across a selected set of genetic variants shared between a discovery sample (some CAD GWAS for example) and target sample (some other genotyped cohort such as the Alzheimerʼs Disease Neuroimaging Initiative, ADNI), resulting in an aggregate score that reflects the genetic contribution to the disease phenotype in the target cohort. AD-PRS have effectively discriminated between AD cases and controls^17^, been used as a predictor of conversion from MCI to AD^18^, been linked to inflammatory biomarkers^19^, CSF amyloid beta levels^20^, CSF tau levels^21^, hippocampal volume^22^, cortical thickness^23^ and age of onset of AD^24^. AD-PRS related work has been recently reviewed by^25^

Just as research has been devoted to investigating how AD-PRS affects AD phenotypes, the same concept can be used to investigate genetic risk for other diseases on AD pathology. Here, we specifically investigate the role of common, small-effect, cardiac-related genetic variants to Alzheimer’s disease. This enables us to elucidate the role that cardiovascular health plays in relation to dementia. While we focus here on the influence of underlying genetics upon WMHs, whole brain atrophy and the changes therein, the cardiac-cerebrovascular axis is no doubt complex, encompassing many biological pathways. We believe, however, that a combination of genetics and imaging, in particular longitudinal images that capture changes over a disease trajectory, will provide important insights into this system.

## 2. Material and methods

Data used in the preparation of this article were obtained from the Alzheimer’s Disease Neuroimaging Initiative (ADNI) database (adni.loni.usc.edu). The ADNI was launched in 2003 as a public-private partnership, led by Principal Investigator Michael W. Weiner, MD. The primary goal of ADNI has been to test whether serial magnetic resonance imaging (MRI), positron emission tomography (PET), other biological markers, and clinical and neuropsychological assessment can be combined to measure the progression of mild cognitive impairment (MCI) and early Alzheimer’s disease (AD). For up-to-date information, see www.adni-info.org. The R-data-package ADNIMERGE (dated 2020-05-19) was used to access ADNI data.

### 2.1 Computing white matter hyperintensities

Regional and total WMH values were determined using the Bayesian Model Selection (BaMoS) software^26^, a white matter lesion segmentation algorithm. BaMoS was applied to 932 ADNIGO and ADNI2 participants following^27^ and described therein. In short, the label fusion algorithm GIF (Geodesic Information Flows)^28^ was used to parcellate the T1-weighted cortical grey matter into various cortical and subcortical brain structures in an automated fashion. It additionally carries out skull stripping and generates probabilistic atlases for each individual. These atlases are then input to BaMoS alongside co-registered FLAIR images consisting of log-transformed normalised intensities. To determine white matter lesions, BaMoS computes the most suitable model description of the data accounting for prevailing outliers. It manages this by first partitioning data into inlier and outlier portions and then modelling input data in a hierarchical fashion, with elements then separated into one of four tissue types – grey matter, white matter, cerebrospinal fluid and non-brain. Each of these in turn are modelled via a Gaussian mixture model with the number of constituent parts determined using a split-and-merge strategy. An Expectation-Maximization (EM) algorithm is used for optimisation with model selection implemented using Bayesian Information Criterion to produce probabilistic lesion maps from which measurements of lesion volumes are inferred. Regional volumes in cubic mm were also computed across five lobes and four radial layers.

### 2.2 Brain atrophy

To compute changes in brain volume we utilise the Boundary Shift Integral (BSI) method^29,30^. Briefly, the BSI is defined as the difference in brain volume (either in total brain or in a brain region via displacement of the boundaries), automatically computed (brain mask creation is semi-automated) between a baseline scan and a repeat scan at a later time. The KN-BSI^30^ is an extension of the classic-BSI and carries out tissue-specific intensity normalisation which deals with tissue-contrast differences, producing a smaller standard deviation in atrophy changes than the classic-BSI. BSI values were obtained from the *foxlabbsi* table in ADNI comprising 2348 subjects (across ADNI1, ADNI2, ADNI3 and ADNIGO), where all T1-weighted scans included in the core datasets pertaining to BSI were obtained using 3T scanners. Some months had very few available scans so these were removed to maintain consistency between scan interval times. MRI scans were originally made using accelerated and non-accelerated acquisitions and we have chosen to focus on the accelerated scans^31^ as they have been shown to result in fewer motion artifacts in pairs of scans resulting from patient movement. Finally, some scans had BSI-determined brain volume increases which may be a result of better subject hydration or due to overall noise. Scans with BSI<0 (corresponding to an increase in whole brain volume over time) were removed from the analysis.

### 2.3 ADNI genetic target data pre-processing

The genetic data used in this work is a combination ADNI1, ADNIGO and ADNI2 participant genotypes, comprising a total of 1674 subjects^32^. Details on quality control and imputation (use of the HRC reference panel, the Sanger server, EAGLE2 for phasing, and PBWT for imputation) are described in^33^. Briefly, following imputation, multi-allelic SNPs and SNPs with INFO score less than 0.3 were removed; then calls with less than 90% posterior probability of the imputed genotype were set to missing. SNPs missing in more than 10% subjects, deviating from Hardy-Weinberg equilibrium (p < 5e-7) and with minor allele frequency less than 5% were all removed. These processing steps were carried out using PLINK v1.9^34^[www.cog-genomics.org/plink/1.9/] and resulted in a final set of 5,082,879 autosomal SNPs. For ancestry determination and relatedness analysis, following^21,33^ a HapMap 3 reference panel was utilised where individuals with greater than 80% Central European ancestry were held. PLINK v1.9 was then used to retain common SNPs with MAF ≥ 5% and carry out LD-pruning and construct a genetic relatedness matrix (threshold=0.1) and filter to remove related subjects. This resulted in the exclusion of 116 subjects most likely due to them being genetically non-central European (there are known to be a number of erroneous self-identifications of European descent in this cohort) or being related to other subjects.

For the CSF amyloid beta measurements we used ADNIMERGE *ABETA.bl* values and removed subjects with missing data and set values recorded as <1700 to 1700 (PET CSF Ab1-42; 192 pg/mL cut-off value; Luminex assay; data range 203 to 1700 with mean=1024.7).

### 2.4 Coronary Artery Disease & Alzheimer’s Disease discovery GWASs

To investigate the PRS contribution due to AD we utilised the largest currently available meta-GWAS of AD featuring 35,274 clinical and autopsy-documented AD cases and 59,163 controls^35^. Summary statistics were downloaded from The National Institute on Aging Genetics of Alzheimerʼs Disease Data Storage Site (NIAGADS; July 2020) comprising 11,480,633 SNPs.

In order to investigate the PRS contribution due to CAD we use a meta-analysis of 60,801 CAD cases and 123,504 controls^36^. Summary statistics were downloaded from the CARDIoGRAMplusC4D (Coronary ARtery DIsease Genome-wide Replication and Meta-analysis (CARDIoGRAM) plus The Coronary Artery Disease (C4D) Genetics) consortium http://www.cardiogramplusc4d.org/data-downloads/ in July 2020 comprising 8,624,384 variants.

Given the very large effect size of APOE-e4 upon AD pathology, we removed the APOE region so as to explore genetic effects beyond this risk factor. The block removed from chromosome 19 (hg19 coordinates) comprises SNPs 44 400 375 (rs430308) to 46 500 052 (rs62113435).

### 2.5 Polygenic Risk Scores

The Polygenic Risk Score (PRS) is a weighted sum of allele counts where the weights are odds ratios (effect sizes) from a discovery GWAS and represent the strength between the variant and the trait it is associated with. Polygenic Risk Scores were computed using PRSice v2.1.9^37^. To ensure that loci are isolated and independent, LD clumping was applied so that SNPs in LD with one another are removed such that the SNP with the lowest p-value within each LD block of correlated SNPs is held for analysis. LD clumping was conducted with clumping window of 250kb on either side of the index SNP, with an r^2^ threshold of 0.1 and p-value threshold of 1.

The PRS for each subject was computed using the “--score avg” setting in PRSice by summing up the product of each variant by the number of effective alleles observed divided by the number of alleles included for that individual. This last divisor makes the PRS scores more comparable between subjects in the presence of missing SNPs. PRS was calculated for each subject for two p-value cutoffs p=1e-5 (i.e., genome-wide suggestive loci) capturing oligogenic effects and p=0.5 capturing polygenic effects, which have been identified as sufficient to encompass threshold-variation^17^.

Of note, it is imperative when working with PRS that there is no sample overlap between discovery GWAS (here Kunkle et al. 2019) and the target cohort (here the combined ADNI1, ADNIGO and ADNI2). We did not encounter any sample overlap as the discovery GWAS use The Alzheimerʼs Disease Genetics Consortium (ADGC) summary statistics, which only use HC and AD cases from ADNI1 comprising of 1.5T scans. However, from the 846 individuals with imaging and genetic data, we excluded 116 subjects with non-central European ancestry, resulting in the final sample size of 730 for our study. Moreover, demographic genetic differences arising from ancestral population structure have been found to bias PRS scores^38^. This is the case particularly for high p-value cut-offs (e.g., p=0.5) leading to PRS with many 1000s of SNPs. To account for this effect our linear regression models include the first five principal components of population structure as covariates.

### 2.6 Second-order grey matter changes

By plotting longitudinal whole brain volume change, or BSI (from baseline in units of cubic cm or ml where 1cm^3^=1ml), for each subject taken at three, six, twelve and twenty-four months post baseline, the gradient of the line of best fit gives a second order change (the second derivative) in whole brain volume, or the rate of change of BSI per subject. We term this rate of change the “atrophy rate”, which provides a measure of how fast the whole brain volume is declining. By way of illustration, if, for example, there is a BSI of 6ml between months 0-24 then the gradient would be 0.25ml/month^2^ (which equals 3ml/yr^2^) and would have an intercept of zero.

### 2.7 Statistical analysis

We first investigated polygenic risk scores for AD (AD PRS) as well as polygenic risk scores for CAD (CAD PRS) and their relationship with total whole brain volume WBV over a range of timepoints (which are normalised by dividing by Intracranial Volume, ICV), total White Matter Hyperintensities WMH (tot) at baseline (which are log-transformed) and CSF amyloid beta measurements. We used linear regressions within each diagnostic group adjusting for age, sex, education, APOE-e4 burden and the first five principal components of population structure. In the case of the WMH (tot) we also included the ICV as an additional covariate. Regressions provided t-values as well as p-values for the association between PRS and biomarkers; given the large number of comparisons (36 = 2 PRS thresholds x 2 PRS x 3 diagnoses x 3 biomarkers) we have adjusted p-values for multiple comparisons using an FDR-correction (rate=5%). Given that both increasing whole brain atrophy, WMHs as well as BSI values are all known to lead to worse health outcomes all p-values are based on one-tailed tests. For the regional WMH data we carried out an identical analysis but over five lobar regions: frontal, parietal, occipital, temporal and a combined basal ganglia, thalami and infratentorial volume. Each of these regions was normalised by their corresponding regional volume. Finally, the occipital region was explored in more detail through four radial layers of increasing distance from the ventricular system, such that the first two layers represent the inner periventricular WMH loads and the final two layers the deep WMH loads.

In addition, for the *atrophy accelerations* we also used linear mixed effects models (with subject as random intercept and time-since-baseline as random slope across diagnostic groups; R library lme4) across both p-value PRS-thresholds. In addition, to explore further how *atrophy accelerations* are associated with polygenic risk we also utilised linear mixed effects models. Here subject is the random effect so the atrophy acceleration is calculated per subject. In this way we allow for a positive correlation among measurements for the same individual once the fixed effects have been accounted for. Time or months since baseline is also included as a random effect. For both of these variables there is one intercept. In order to get the influence of PRS on the ʻrate of atrophyʼ (atrophy acceleration) the model includes the PRS-by-time interaction (*PRS*time*). This allows different PRSs to influence the rate of BSIs. This is all implemented using the R library function lme4 with dependent variable the BSI measurement and the independent variable *PRS*time* or polygenic score-by-time interaction, where time is the months since baseline) across both p-value PRS-thresholds:

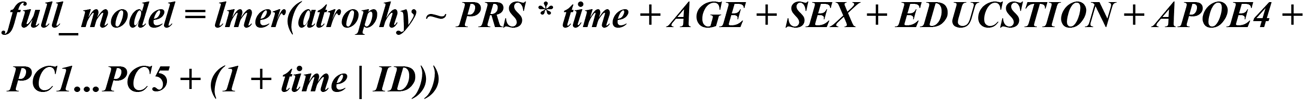

Similar covariates as used in previous linear regressions and with the *PRS*time* comprise the fixed effects. We used a likelihood ratio test to compare full models and reduced models (without the polygenic score-by-time interaction in the reduced model) to compute model p-values:

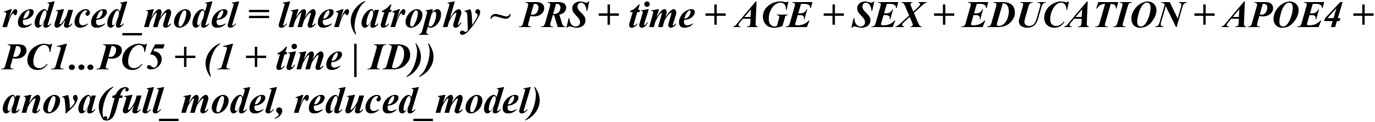

We used a likelihood ratio test to compare full models and reduced models (without the polygenic score-by-time interaction) to compute model p-values.

## 3. Results

### 3.1 Filtering core datasets

A total of 2269 unique subjects were extracted from ADNIMERGE of which 2257 had available baseline diagnoses. To maximise the numbers per diagnostic group, sub-diagnostic groups were merged: (1) ˭CNˮ (Control/Normal) and ˭SMCˮ (Significant Memory Concern) to ˭HCˮ (Healthy Control); (2) ˭EMCIˮ (Early Mild Cognitive Impairment) and ˭LMCIˮ (Late Mild Cognitive Impairment) to Mild Cognitive Impairment label ˭MCIˮ; (3) ˭ADˮ (Alzheimer’s Disease) remaining unchanged. A subset of these subjects with computed total and regional WMH using BaMoS (see below) results in a cohort of 871 individuals. Further subjects were excluded following addition of AD PRS and later CAD PRS (134,456 and 135,584 variants respectively) and principal components of population structure, leaving N=730 subjects that comprise CORE DATASET 1 (*Figure 1*).

**Figure 1:**
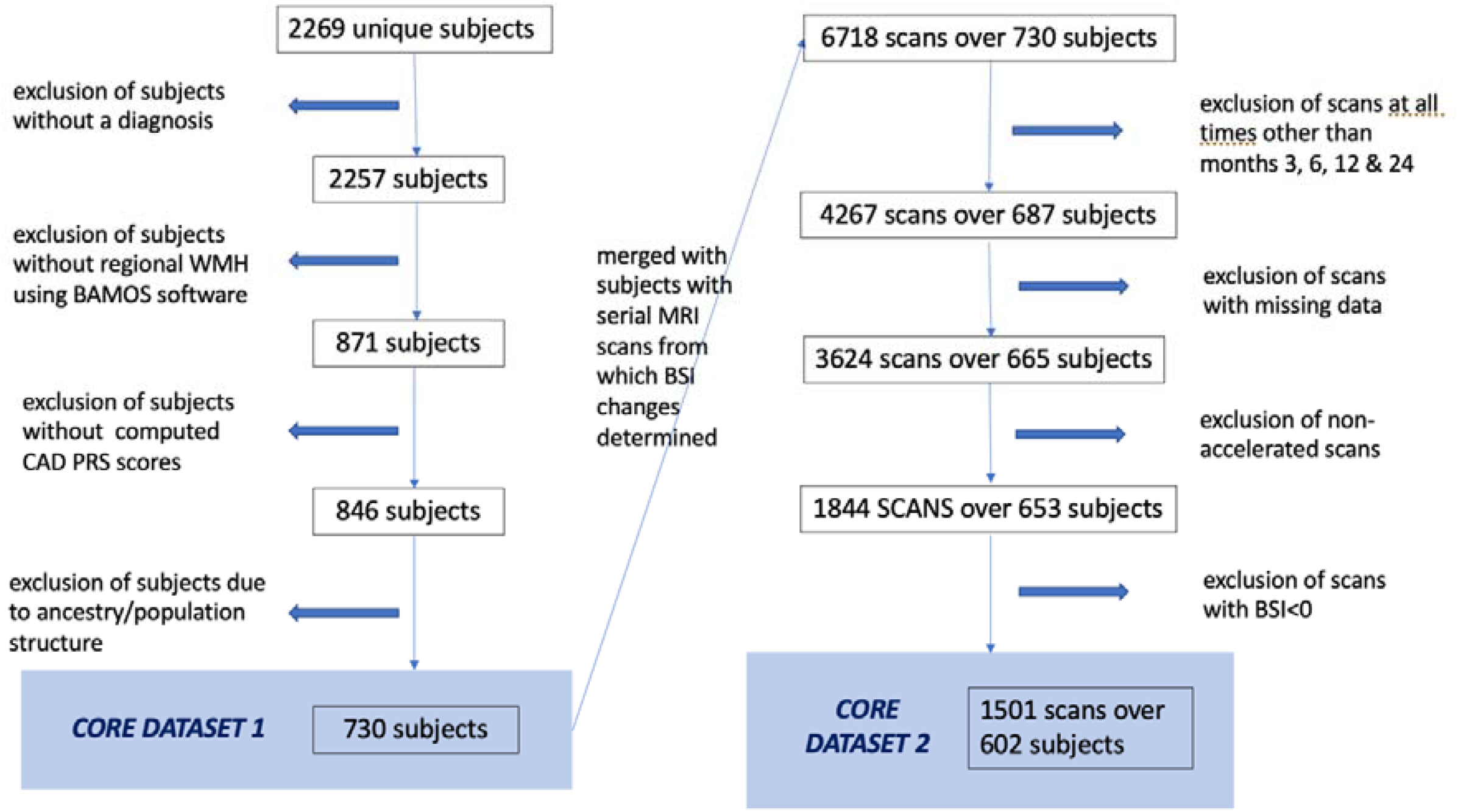
Illustration of the selection of subjects filtered for analysis leaving 730 subjects that comprise CORE DATASET 1 and 602 subjects that comprise CORE DATASET 2.

We augmented CORE DATASET 1 with available boundary shift integral (BSI) data (ADNIMERGE *foxlabbsi* table) to investigate atrophy accelerations, i.e., changes in whole brain volumes over time. Given the paucity of scans at months 8, 18, 36, 48, 60, 72, 84, 96, 120, 132, 144 and 156 (see supplementary *Figure S2*) only scans from months 3, 6, 12 and 24 were retained (*Figure S3*). The remaining atrophy values comprised 824 instances of negative total KN-BSIs (across all months), implying a growth in whole brain volume over time. As this is unlikely these scans were considered to be inaccurate and removed from the sample which resulted in a loss of 51 subjects. This formed CORE DATASET 2 which comprises 1501 scans over N=602 subjects (*Figure 1*).

#### 3.1.1 Core dataset summaries

*Table 1* shows the demographics of CORE DATASET 1 and 2 respectively. There were significant differences in age and years of education between the diagnostic categories *Table S1* (and *Figure S3*) show the number of follow-up MRI scans available within each diagnostic group. The proportions match the sample-size distributions. However, only a few scans were available at the 24-month mark for the AD subgroup.

**Table 1:** Baseline demographics of Table 1a (top) CORE DATASET 1 and Table 1b (bottom) CORE DATASET 2. For CORE DATASET 1 more than half of the subjects fall in the MCI diagnostic category, almost a third HC and less than a fifth AD. The HC group has a higher fraction of females and as expected the AD participants are slightly older on average and there is a slight decrease in time spent in education for the AD subset (one-way ANOVA - age: p=1.49e-05; education: p= 0.0064). In CORE DATASET 2 we see a similar breakdown of sample sizes, sex, age and education by diagnostic group one-way ANOVA - age: p=2.51e-05; education: p=0.06).

|  | HC | MCI | AD | TOTAL |
| --- | --- | --- | --- | --- |
| N | 218 (29.9%) | 390 (53.4%) | 122 (16.7%) | 730 |
| Female (%) | 117(53.7%) | 171 (43.8%) | 51(41.8%) | 339 (46.4%) |
| Age (SD) | 73.4 (5.9) | 71.5 (7.5) | 74.8 (8.0) | 72.6 (7.2) |
| Education (SD) | 16.7 (2.5) | 16.3 (2.6) | 15.7 (2.7) | 16.3 (2.6) |
| N | 172 (28.6%) | 336 (55.8%) | 94 (15.6%) | 602 |
| Female (%) | 91 (52.9%) | 151 (44.9%) | 37 (39.4%) | 279 (46.3%) |
| Age (SD) | 73.7 (6.1) | 71.7 (7.3) | 75.1 (7.5) | 72.8 (7.1) |
| Education (SD) | 16.6 (2.5) | 16.4 (2.6) | 15.8 (2.7) | 16.3 (2.6) |

### 3.2 White matter lesions

WMH volume was greater in the AD group (Tukey multiple comparisons of means, AD-to-MCI:p_adj_=0.007, AD-to-HC:p_adj_=0.03) and marginally greater in the MCI group over the healthy controls (*Figure S1, a relationship which holds when WMH is corrected for subject age;* WMH volume was also greater in the AD group when broken down by sex; Table S2). Following an exploration of regional WMH (including total frontal, parietal, temporal and occipital WMH volumes (results not shown) the most compelling result was the combined basal ganglia and infratentorial WMH volumes in male subjects, which showed a clear difference between diagnostic groups (*Figure S4*).

### 3.3 Polygenic risk

Our linear regression analysis for an effect between AD PRS or CAD PRS (over both PRS thresholds) on baseline GM, total WMH or CSF amyloid did not find any statistically significant results (P_FDR_>0.05; *Table 2*). However, for individuals with an AD diagnosis both AD PRS (P_FDR_=0.03) and CAD PRS (P_FDR_=0.04) are associated with occipital lobe WMH at the PRS-threshold of p=0.5 (*Table 4*).

**Table 2:** Results from CORE DATASET 1 using the low p-cut-off 1e-05 and the high p-cut-off 0.5. t-values (p-values, one-tailed FDR-corrected) to 2sf. following linear regression with confounders for age, sex, education, APOE-e4 burden and first five principal components of population structure (additional ICV confounder for log transformed WMH regression) across diagnostic groups for AD PRS and CAD PRS; WBV normalised by ICV; AD PRS and CAD PRS both exclude APOE region. For amyloid beta, subjects with missing data were removed from the linear model (n=665).

|  |  | HC |  | MCI |  | AD |  |
| --- | --- | --- | --- | --- | --- | --- | --- |
|  |  | AD PRS | CAD PRS | AD PRS | CAD PRS | AD PRS | CAD PRS |
| P=1e-05 | WBV | 0.61 (0.30) | 1.9 (0.27) | -2.2 (0.07) | 1.8 (0.34) | 1.0 (0.49) | -0.78 (0.47) |
|  | WMH (tot) | 0.21 (0.44) | 0.31 (0.39) | -1.08 (0.34) | -1.43 (0.48) | 1.93 (0.09) | -0.78 (0.13) |
|  | Amyloid Beta | -0.89 (0.20) | 2.4 (0.16) | 0.1 (0.48) | 1.1 (0.47) | -0.35 (0.38) | 0.13 (0.45) |
| P=0.5 | WBV | -0.8 (0.30) | 0.85 (0.26) | -0.96 (0.31) | 0.24 (0.42) | 0.31 (0.49) | -0.23 (0.47) |
|  | WMH (tot) | 0.68 (0.36) | 0.48 (0.39) | -0.73 (0.34) | -0.12 (0.48) | 1.63 (0.09) | 1.35 (0.13) |
|  | Amyloid Beta | -0.89 (0.20) | 0.13 (0.48) | -0.46 (0.48) | -0.49 (0.47) | -1.2 (0.20) | 1.8 (0.12) |

**Table 3:** Regional WMH analysis from CORE DATASET 1 using the low p-cut-off 1e-05 and the high p-cut-off 0.5. T-values (p-values, one-tailed FDR-corrected) to 2dp. following linear regression with confounders for age, sex, education, APOE-e4 burden and first five principal components of population structure and natural log of regional WMH normalised by regional volume.

|  |  | HC |  | MCI |  | AD |  |
| --- | --- | --- | --- | --- | --- | --- | --- |
|  |  | AD | CAD | AD | CAD | AD | CAD |
|  |  | PRS | PRS | PRS | PRS | PRS | PRS |
| P=1e-05 | frontal | 0.29<br>(0.45) | 0.24<br>(0.41) | -0.98<br>(0.34) | -2.0<br>(0.21) | 1.8<br>(0.29) | -1.2<br>(0.25) |
|  | parietal | -0.15<br>(0.49) | 0.52<br>(0.45) | -1.0<br>(0.49) | -0.95<br>(0.49) | 1.8<br>(0.12) | -1.5<br>(0.16) |
|  | occipital | 0.66<br>(0.45) | 0.29<br>(0.46) | -1.1<br>(0.17) | -1.6<br>(0.5) | 1.4<br>(0.06) | -0.96<br>(0.2) |
|  | temporal | -0.03<br>(0.44) | 1.2<br>(0.31) | -0.13<br>(0.42) | -1.3<br>(0.45) | 2.3<br>(0.38) | -0.69<br>(0.28) |
|  | basal ganglia +<br>thalami +<br>infratentorial | 0.28<br>(0.46) | -0.15<br>(0.47) | -1.0<br>(0.11) | -0.53<br>(0.46) | 0.93<br>(0.49) | -0.21<br>(0.47) |
| P=0.5 | frontal | 0.64<br>(0.38) | 0.76<br>(0.27) | -0.26<br>(0.34) | -0.10<br>(0.49) | 1.1<br>(0.29) | 0.80<br>(0.25) |
|  | parietal | 0.52<br>(0.49) | 0.29<br>(0.45) | -0.47<br>(0.47) | -0.09<br>(0.49) | 1.5<br>(0.12) | 1.2<br>(0.16) |
|  | occipital | 0.48<br>(0.34) | 0.27<br>(0.46) | -0.92<br>(0.17) | 0.03<br>(0.5) | 1.7<br>(0.03) | 2.0<br>(0.04) |
|  | temporal | 0.53<br>(0.34) | 0.69<br>(0.31) | -1.3<br>(0.20) | -0.31<br>(0.45) | 2.0<br>(0.39) | 1.3<br>(0.17) |
|  | basal ganglia +<br>thalami +<br>infratentorial | 0.71<br>(0.46) | -0.42<br>(0.45) | -0.71<br>(0.05) | 0.15<br>(0.46) | 0.76<br>(0.49) | 0.35<br>(0.47) |

**Table 4:**
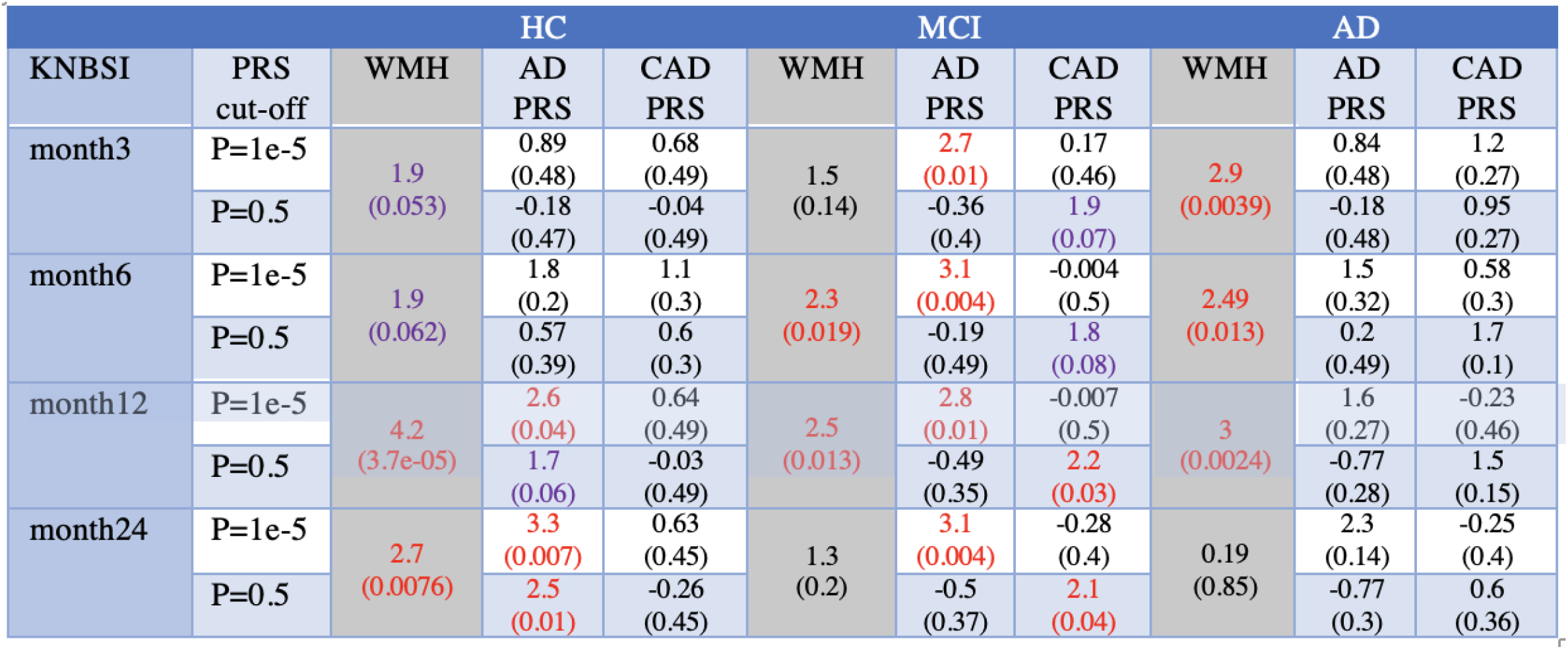
Results from CORE DATASET 2. T-values (p-values) for a range of KN-BSI whole brain atrophy over scans at months 3, 6, 12 and 24 with respect to natural log of WMH (at baseline or month 0), CAD PRS and AD PRS in different diagnostic groups; linear regression with confounders for age, sex, education, APOE-e4 burden and five principal components of population structure (including ICV confounder for WMH regression) across diagnostic groups for two PRS thresholds; APOE region excluded in AD PRS and CAD PRS. All PRS regression p-values are corrected for multiple testing and are one-tailed.

### 3.3 Whole brain atrophy

Given that a snapshot of whole brain volume at baseline may not be sufficiently informative, we investigated the effect of total WMH at baseline and PRS on longitudinal changes in whole brain volumes. Following the baseline scan, KN-BSI increases (representing a decrease in whole brain volume) with passing months in all three diagnostic groups (*Figure 2*), but this is most pronounced in the AD cohort (see *Figure S4* for multiple comparison p-values). Using linear regression, we found a statistically significant effect (P_FDR_<0.05) of baseline WMH on WBV atrophy in all diagnostic groups over differing times (*Table 4*). This association also held in a corresponding regional analysis of frontal, parietal, occipital, temporal and combined basal ganglia, thalami and infratentorial WMH volumes (*Table S3*) with WBV drop being particularly correlated over months 12 and 24 (especially in the MCI cohort).

**Figure2:**
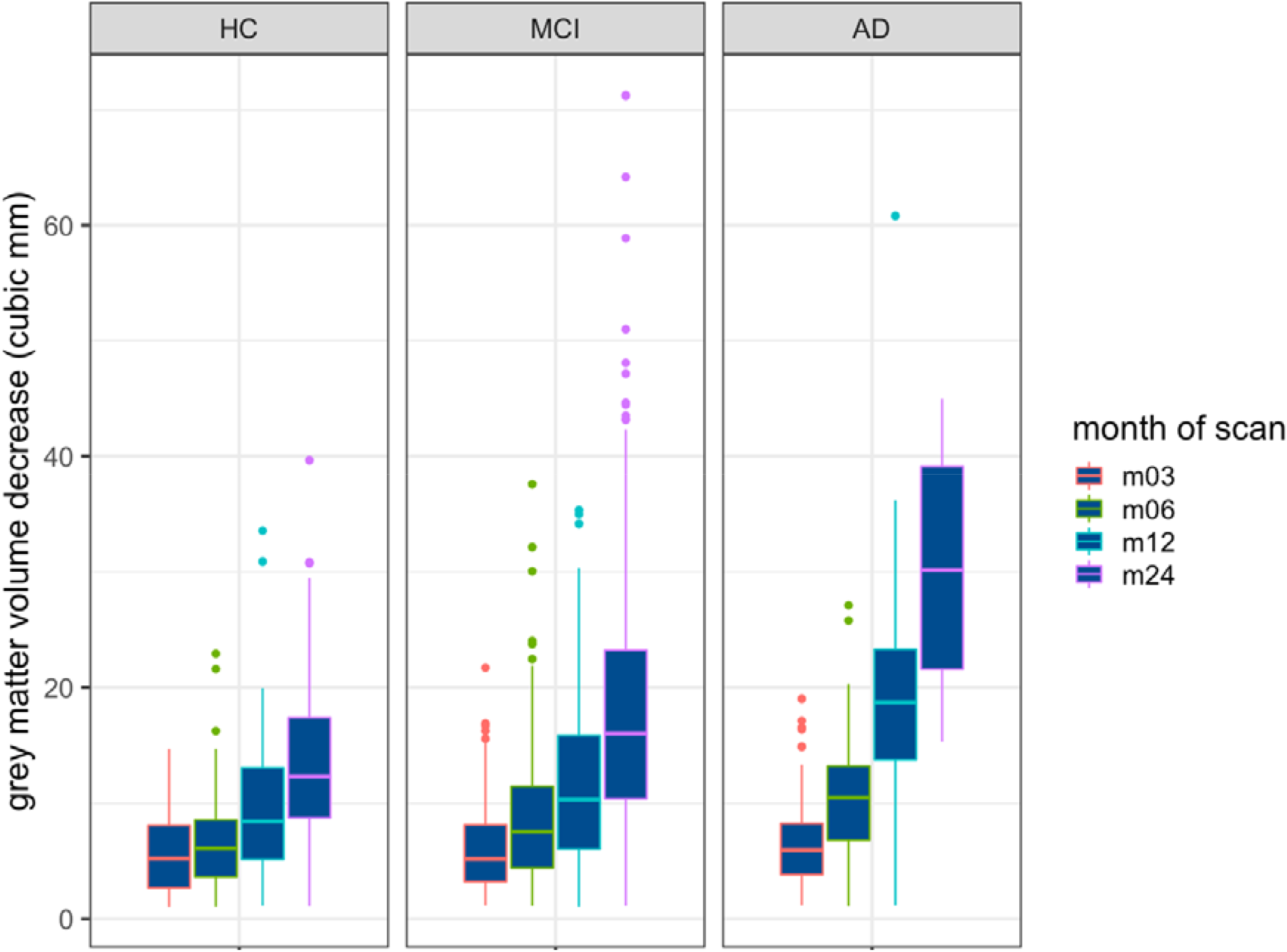
whole brain atrophy (KN-BSI) in each diagnostic group across all time intervals – m03, m06, m12 and m24 represent 3, 6, 12 and 24 months following baseline scan respectively.

AD PRS is associated with whole brain atrophy (KN-BSI) in HC and MCI, but not in subjects with AD. Moreover, in the MCI cohort AD PRS is correlated with brain atrophy at each time-point but only for the p=1e-5 PRS threshold. Finally, in the HC cohort the KN-BSI and AD PRS are correlated in the later month 12 and month 24.

There is no statistically significant correlation between CAD PRS and KN-BSI in both the HC and AD diagnostic groups. However, in the MCI cohort we see a correlation in the later month 12 and month 24, but only for the p=0.5 threshold. So in the p=0.5 threshold for month 12 and month 24, the CAD PRS is more correlated with whole brain atrophy than the AD PRS (month 12: t=2.2, P_FDR_=0.03; month 24: t=2.1, P_FDR_=0.04).

### 3.4 whole brain atrophy acceleration

To probe the *rate of change* of whole brain volume over longitudinal scans per subject with respect to the underlying genetics we enumerate the ‘atrophy acceleration’ in *Table 5a*.

**Table 5:**
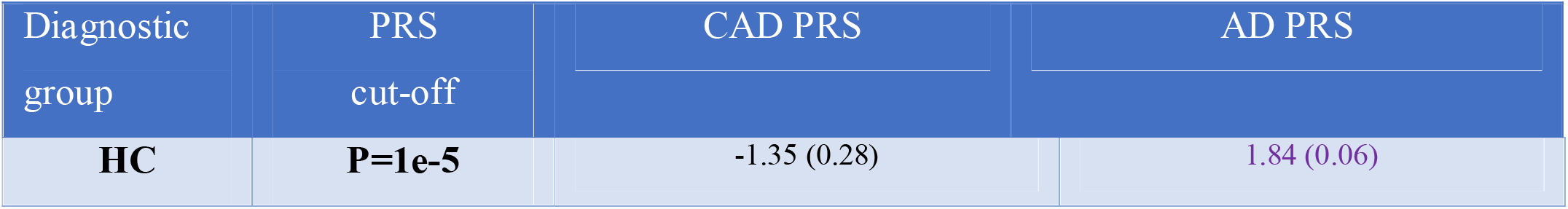
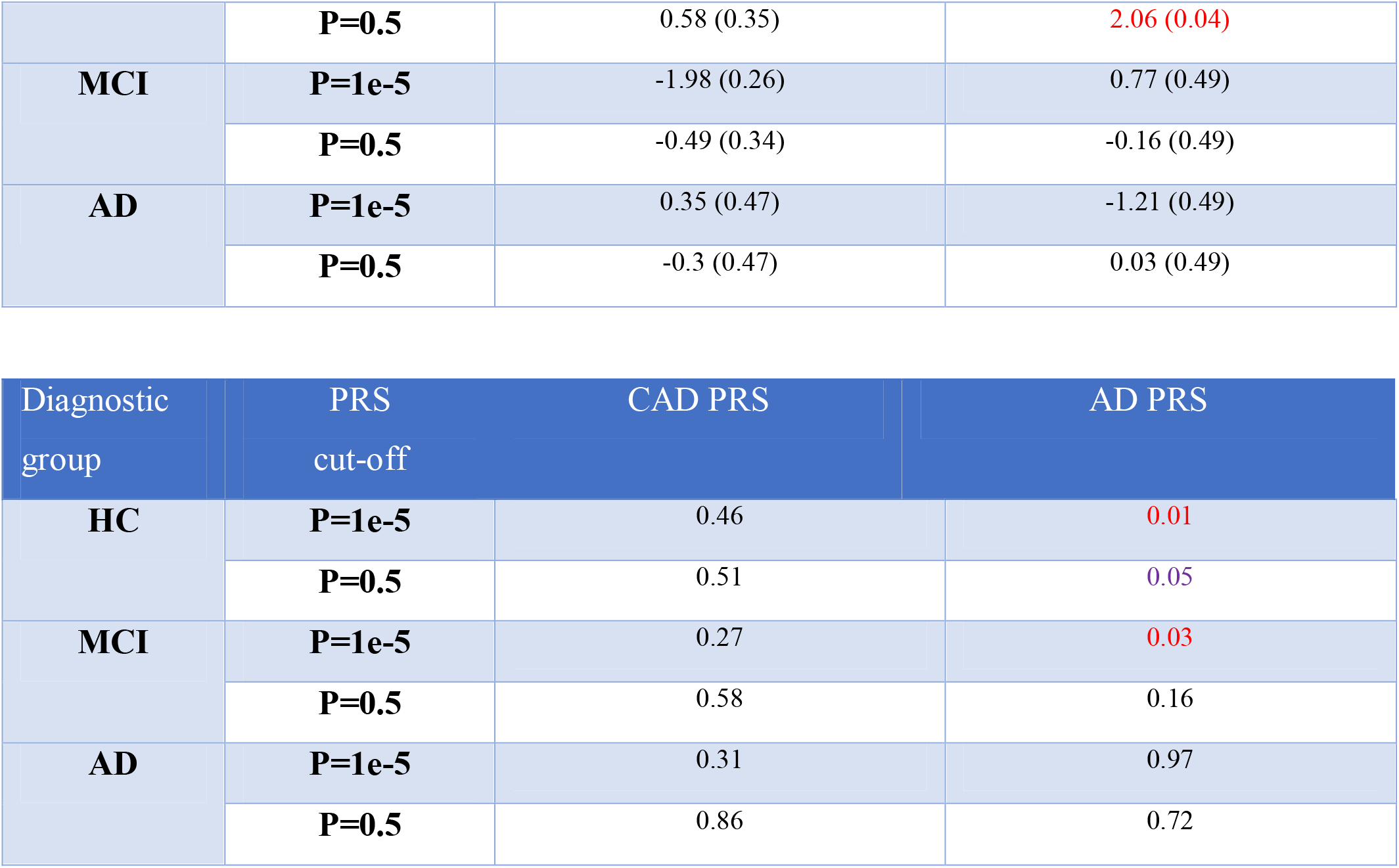
**Table 5a (top) T-values (p-values) for atrophy acceleration, or, rate of change of whole brain volume (gradient from line of best fit over serial scans at months 3, 6, 12 and 24 per subject) with respect to AD PRS and CAD PRS.** Following linear regression with confounders for age, sex, education, APOE-e4 burden and five principal components of population structure across diagnostic groups for two PRS thresholds; APOE region excluded in AD PRS and CAD PRS. All PRS regression p-values are corrected for multiple testing and are one-tailed. **Table 5b (bottom) P-values from linear mixed effects model with subject as random intercept and time-since-baseline as random slope.** Across diagnostic groups and oligogenic and polygenic thresholds for atrophy acceleration or rate of change whole brain volume (gradient from line of best fit over serial scans at months 3, 6, 12 and 24 per subject) with respect to CAD PRS-and AD PRS-by-time interaction.

*Atrophy acceleration* is significantly greater in the AD cohort compared to HC and MCI (p<4.3e-13; *Figure 3*). While there is no correlation between CAD PRS and atrophy acceleration (*Table 5a*), there is a statistically significant correlation between AD PRS and atrophy acceleration for the p=0.5 threshold in the healthy controls (HC). It is also notable that at the p=1e-5 threshold this correlation shows a statistical trend (P_FDR_=0.06). Our mixed effect analysis (*Table 5b*) showed that there is no association between longitudinal changes in grey matter (quantified by the BSI) and time-by-CAD polygenic score interaction, however, time-by-AD polygenic score interaction is correlated with longitudinal WBV changes in both the HC (P_FDR_ =0.01) and MCI (P_FDR_ =0.03) subsets at the p=1e-5 PRS-threshold.

**Figure 3:**
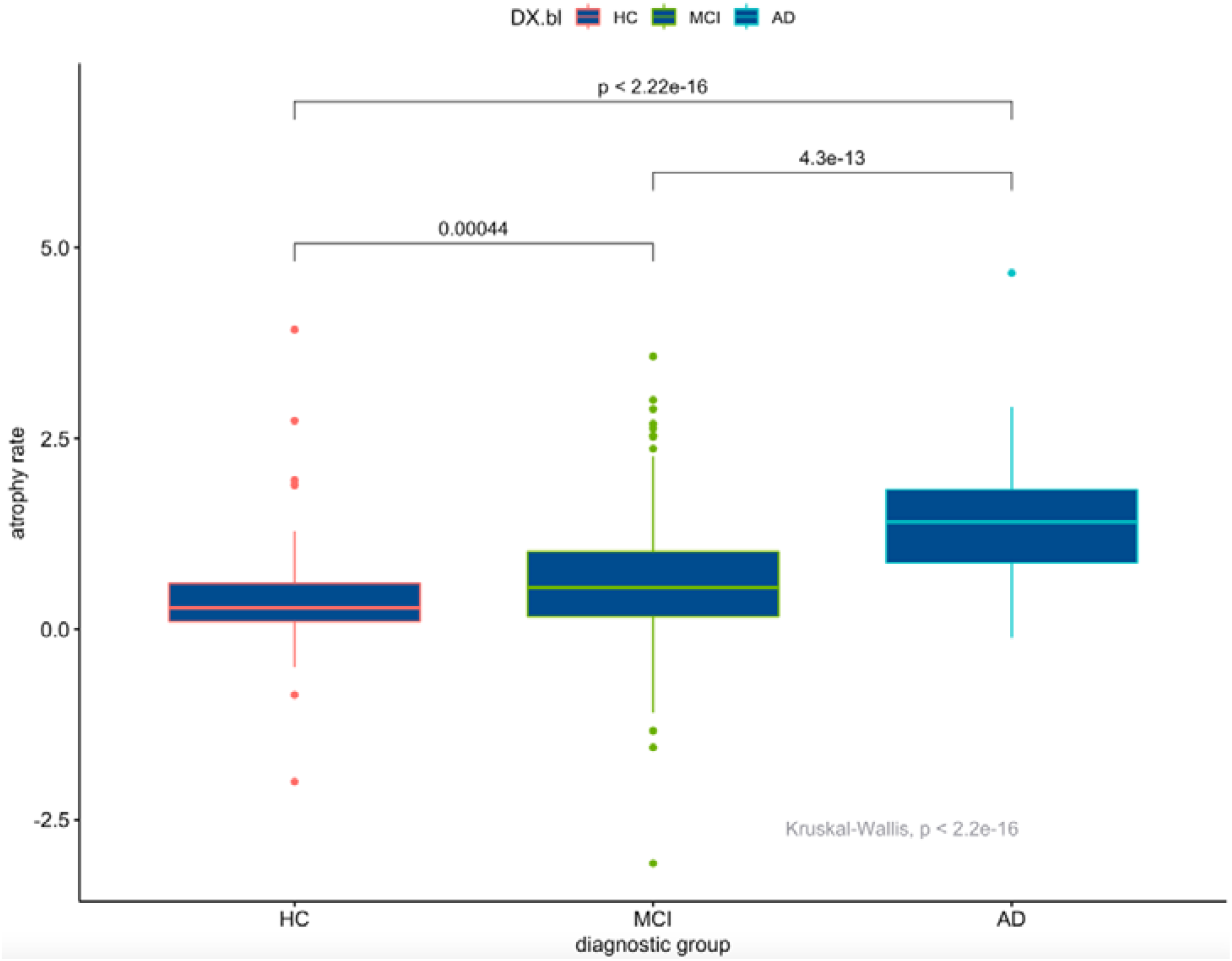
Median atrophy acceleration in cubic cm/month^2 across diagnostic groups. AD subjects whole brain volume is decreasing much faster than MCI, which are decreasing faster than HC. Multiple comparison Kruskal-Wallis p-values shown.

## 4. Discussion

In this study we aimed to investigate possible associations between cardiac genetics and Alzheimer’s dementia. We focused upon the genetic variants outside of the APOE locus that have been shown to be associated with AD, and separately, with CAD genetic variants. In particular we looked at their impacts upon white matter lesions and whole brain volume changes. We found a correlation between coronary artery genetic risk and whole brain atrophy suggesting that many small effect-size variants contribute to neuronal loss in mildly cognitively impaired (MCI) individuals. Surprisingly, under certain PRS-thresholds and at certain times in the disease-course the underlying polymorphisms for coronary artery disease may well have more of an impact than those of AD. We have also highlighted how whole brain atrophy acceleration is associated with Alzheimer’s disease polygenic risk score outside the APOE locus in healthy individuals. Further, we have shown for the first time that genetic variants that contribute to both AD and CAD beyond APOE are a strong predictor of white matter lesions in the occipital lobe in subjects already diagnosed with AD.

White matter lesions were clearly higher in the AD group, something that is driven in part by age (*Figure S5*). While all linear models treated age as a covariate, disentangling white matter lesion increase as a function of time separate to any age-related AD pathologies is challenging. This may well be of great import if early pharmaceutical intervention is necessary to curtail AD in later life as WMH extent could be utilised as a surrogate biomarker for downstream dementia^39^.

No associations were found between AD PRS or CAD PRS and baseline total WBV and WMH volumes across all diagnostic groups, although PRSs have been shown to be predictive of AD risk^40^. That said, AD PRS beyond APOE is correlated with white matter load in the occipital lobe in AD subjects upon the inclusion of many variants (polygenic p=0.5). In fact, our regional analysis of WMH showed statistically significant association between both AD PRS and CAD PRS with occipital lobe WMH lesion volume in the AD group at a PRS-threshold of p=0.5. This confirms the presence of significant WMH lesions in this lobe seen in other dementia cohorts^41^ and the finding that occipital WMH is correlated with reduced executive function^42^. This may also be related to the cerebral amyloid angiopathy pathway. We were unable to establish this white matter load association by occipital lobe layer, however, this is to our knowledge the first confirmation of a genetic link with this regional WMH and a genetic link to coronary artery disease.

There are also no statistically significant correlations with CSF amyloid beta and CAD or AD PRS, presumably we did not find evidence for an association with AD PRS because amyloid biomarkers are mainly influenced by the APOE-e4 genotype^21^. Moreover, the analysis was conducted within disease groups and not between-disease-groups where stronger differences are known to exist^43^.

When it came to investigating whole brain changes over time our analysis confirmed an association of baseline WMH with WBV decline across all three diagnostic groups. WMH here was derived from scans at baseline and not at the time of repeat scans, thus, WMH burden may indeed serve as an indicator of near-term or future decreases in whole brain volume. The strength of these associations was driven by sample size and time between baseline and follow-up WBV measure. These results confirm earlier studies^41,44^ that showed higher white matter lesion load is correlated with decreasing whole brain volume, albeit using cross-sectional WBV measures. Earlier studies investigating longitudinal whole brain volume change only found an association with WMH in healthy controls ^45^ or hippocampal atrophy in MCI subjects^46^, in contrast to our analysis which extends this correlation to participants with AD.

In the matter of whether PRS are predictive of longitudinal decline in WBV, our analysis showed correlations between AD PRS and WBV decline in the HC group (later months) and MCI group (all months), but not the AD group. Decline in the healthy controls group reflects regional WBV decline with respect to AD PRS observed previously^47^. The lack of a polygenic effect in the AD group, may originate from either limited statistical power, the overall extent of WBV damage in AD or that while AD is advanced WBV volume decrease is driven by other factors beyond AD-risk variants. As in the HC group, the AD PRS, like baseline WMH, is correlated with WBV decrease in later months 12 and 24, which raises the question as to whether genetic effects become more influential with time. Before this can be answered we would have to account for there being more samples in months 12 and 24, and the fact that picking up changes in WBV volumes over months 3 and 6 will be more difficult as they will likely be more subtle. Interestingly, the AD PRS correlation with WBV atrophy in the MCI group was significant at all time points for the oligogenic (p=1e-5) PRS-threshold but not the polygenic (p=0.5) PRS-threshold. This suggests that there are some larger effect-size, common variants (i.e., the peaks in the Manhattan plot outside the *APOE* locus) that are predictive of brain atrophy in persons that already display some cognitive impairment. This result confirms the hypothesis that a small number of AD-SNPs allow maximal predictive power in AD-related subjects^48^. MCI, however, is a heterogeneous group of which only a subset exhibit MCI due to an underlying AD pathology. This is also the most bimodal group in terms of whether they have AD (progressors vs. not progressors). Thus, the AD PRS may correctly predict WBV decline in subjects who have MCI due to AD.

The motivation for this study was an exploration of the genetic effects of coronary artery disease on Alzheimer’s disease. The CAD PRS was only correlated with WBV decline in the MCI group reflecting incidence-based relationships seen in other studies^49^. Again, the PRS-WBV atrophy association was strongest in the later months 12 and 24 but showed a statistical trend in the earlier months 3 and 6. In the MCI group, CAD PRS is significantly correlated with WBV atrophy for polygenic effects (p=0.5 threshold) but AD PRS is significantly correlated with WBV atrophy for oligogenic effects (p=1e-5 threshold). One interpretation might be that the more genetic variants included, the more important the role of cardiac genetics over AD genetics in individuals with mild cognitive impairment. Another interpretation may be that AD PRS is contributing to individuals with MCI due to developing AD dementia, whereas CAD PRS is bestowing subjects a more vascular component to their cognitive loss.

Recent work^50,51^ investigating the genetic architecture of AD with regards to PRS thresholds argues that the polygenic threshold is optimal. Their work shows that studies using an oligogenic threshold ignore the fact that there will be fewer APOE-e4 carrying individuals in the older category biasing results against high p-value threshold variants. In our study we removed the APOE locus so as to explore genetic changes outside this genetic region, however, all linear regressions include APOE-e4 burden as a confounding variable. Investigators have shown that APOE’s effect on AD is greater in older cohorts and suggest that variants outside of APOE could contribute to AD in older persons^52^. As APOE has been considered a target for both the treatment of coronary heart disease and AD these two pathologies may both be affected by similar pathways. We examined regression models with and without the APOE-e4 covariate using ANOVA. The inclusion of the APOE-e4 burden improved all models significantly (p_adj_ < 0.01) with the exception being WBV volume decline in months 3 (p_adj_=0.8) and 6 (p_adj_=0.1) in the HC group (results not shown).

Brain volume is decreasing faster in AD subjects than MCI subjects, where in turn it is decreasing faster than in healthy individuals (Figure 3). The study of polygenic risk suggests that many small effect-size AD variants beyond APOE are a predictor of atrophy acceleration in healthy individuals (and perhaps also MCI subjects). This is in many ways a surprising result as one would expect the AD cohort (and perhaps the MCI cohort) to be comprised of persons whose WBV decrease is accelerating as the pathology develops. Some caution is warranted here given that AD cohort subjects have fewer longitudinal scans thereby biasing the computed gradient. That said, the HC correlation may indicate that genetic effects modulate the speed at which brain cells atrophy during healthy stages. What we do not know is whether a larger atrophy acceleration while healthy makes AD inevitable, more likely or in no way indicative of downstream pathology. However, it is reassuring that the AD PRS in healthy controls is corroborated in the mixed effects models, which are more flexible, embodying both random intercepts and random slopes so as to more realistically capture the subject-level heterogeneity.

The BSI data available through ADNI are not mid-point symmetric: WBV change going from scan A-to-C is not the same as adding WBV volume changes in scan A-to-B and scan B-to-C. The implication is that one has to be cautious about saying anything about the time-varying nature of these non-symmetric BSIs. We investigated this effect using symmetric BSI data ^53^ for the same ADNIGO and ADNI2 subjects (n=572; 3T, accelerated, identical scanner protocol, removing BSI<0, only months 3, 6, 12 and 24 post baseline). Repeated analysis on this symmetric BSI data resulted in the same statistically significant outcomes as described in this work (results not shown). The only exception to this is that the correlation between baseline WMH and WBV atrophy in the HC group was no longer replicated. We are also cautious about over-interpreting our results: while the BSI does measure changes at the border of the brain, it does not mean that the change has actually occurred there. Tissue may have been lost from the middle of the white matter with this change being measured at the edge of the brain.

Given that both whole brain atrophy and whole brain atrophy acceleration are both shown to be correlated with polygenic scores it may be that utilising such genetic summary information (perhaps alongside other routinely collected health measures) can one day be used at birth (or middle age) as a predictor late-life cognitive problems^54^. If so, and if such risk is driven in part by cardiac health, it may be that this risk can be reduced through wellness and behaviour changes in, early to mid-life^55^. Of course, this does depend on whether healthy individuals with such brain changes go on to develop dementia, something only larger longitudinal studies over many years can answer.

## 5. Conclusions

The link between CAD and AD is gaining more attention with one recent study identifying 23 brain regions associated with both cardiovascular disease and Alzheimer’s dementia^56^. WMH loads have been linked to hypertension, hypercholesterolemia and BMI in middle-aged subjects, all of which also contribute to cardiac health^57^. While our work offers some support for the importance of genetic variants known to effect coronary heart disease also being involved in cognitive decline in the elderly, more data is needed to validate these findings. Similarly, the usefulness of WMHs and atrophy accelerations need to be investigated further. In particular serial WMH measures alongside WBV changes would be a more realistic guide to cerebral changes. Further understanding of this pathway from gene to brain will come from cardiac imaging of the heart linked to CAD genetics alongside routinely measured biomarkers over the life-course of many individuals such as those found in large scale, data-rich, longitudinal cohort studies.

## Data Availability

All data produced in the present study are available upon reasonable request to the authors

## Acknowledgements

This research was supported by the National Institute for Health Research University College London Hospitals Biomedical Research Centre. Andre Altmann holds an MRC eMedLab Medical Bioinformatics Career Development Fellowship. This work was supported by the Medical Research Council [grant number MR/L016311/1].

Data collection and sharing for this project was funded by the Alzheimerʼs Disease Neuroimaging Initiative (ADNI) (National Institutes of Health Grant U01 AG024904) and DOD ADNI (Department of Defense award number W81XWH-12-2-0012). ADNI is funded by the National Institute on Aging, the National Institute of Biomedical Imaging and Bioengineering, and through generous contributions from the following: AbbVie, Alzheimer’s Association; Alzheimer’s Drug Discovery Foundation; Araclon Biotech; BioClinica, Inc.; Biogen; Bristol-Myers Squibb Company; CereSpir, Inc.; Cogstate; Eisai Inc.; Elan Pharmaceuticals, Inc.; Eli Lilly and Company; EuroImmun; F. Hoffmann-La Roche Ltd and its affiliated company Genentech, Inc.; Fujirebio; GE Healthcare; IXICO Ltd.; Janssen Alzheimer Immunotherapy Research & Development, LLC.; Johnson & Johnson Pharmaceutical Research & Development LLC.; Lumosity; Lundbeck; Merck & Co., Inc.; Meso Scale Diagnostics, LLC.; NeuroRx Research; Neurotrack Technologies; Novartis Pharmaceuticals Corporation; Pfizer Inc.; Piramal Imaging; Servier; Takeda Pharmaceutical Company; and Transition Therapeutics. The Canadian Institutes of Health Research is providing funds to support ADNI clinical sites in Canada. Private sector contributions are facilitated by the Foundation for the National Institutes of Health (www.fnih.org). The grantee organization is the Northern California Institute for Research and Education, and the study is coordinated by the Alzheimer’s Therapeutic Research Institute at the University of Southern California. ADNI data are disseminated by the Laboratory for Neuro Imaging at the University of Southern California.

We thank the International Genomics of Alzheimerʼs Project (IGAP) for providing summary results data for these analyses. The investigators within IGAP contributed to the design and implementation of IGAP and/or provided data but did not participate in analysis or writing of this report. IGAP was made possible by the generous participation of the control subjects, the patients, and their families. The i–Select chips was funded by the French National Foundation on Alzheimerʼs disease and related disorders. EADI was supported by the LABEX (laboratory of excellence program investment for the future) DISTALZ grant, Inserm, Institut Pasteur de Lille, Université de Lille 2 and the Lille University Hospital. GERAD/PERADES was supported by the Medical Research Council (Grant n° 503480), Alzheimerʼs Research UK (Grant n° 503176), the Wellcome Trust (Grant n° 082604/2/07/Z) and German Federal Ministry of Education and Research (BMBF): Competence Network Dementia (CND) grant n° 01GI0102, 01GI0711, 01GI0420. CHARGE was partly supported by the NIH/NIA grant R01 AG033193 and the NIA AG081220 and AGES contract N01–AG–12100, the NHLBI grant R01 HL105756, the Icelandic Heart Association, and the Erasmus Medical Center and Erasmus University. ADGC was supported by the NIH/NIA grants: U01 AG032984, U24 AG021886, U01 AG016976, and the Alzheimerʼs Association grant ADGC–10–196728.

