## Supplementary material for "Polygenic coronary artery disease association with brain atrophy in the cognitively impaired"

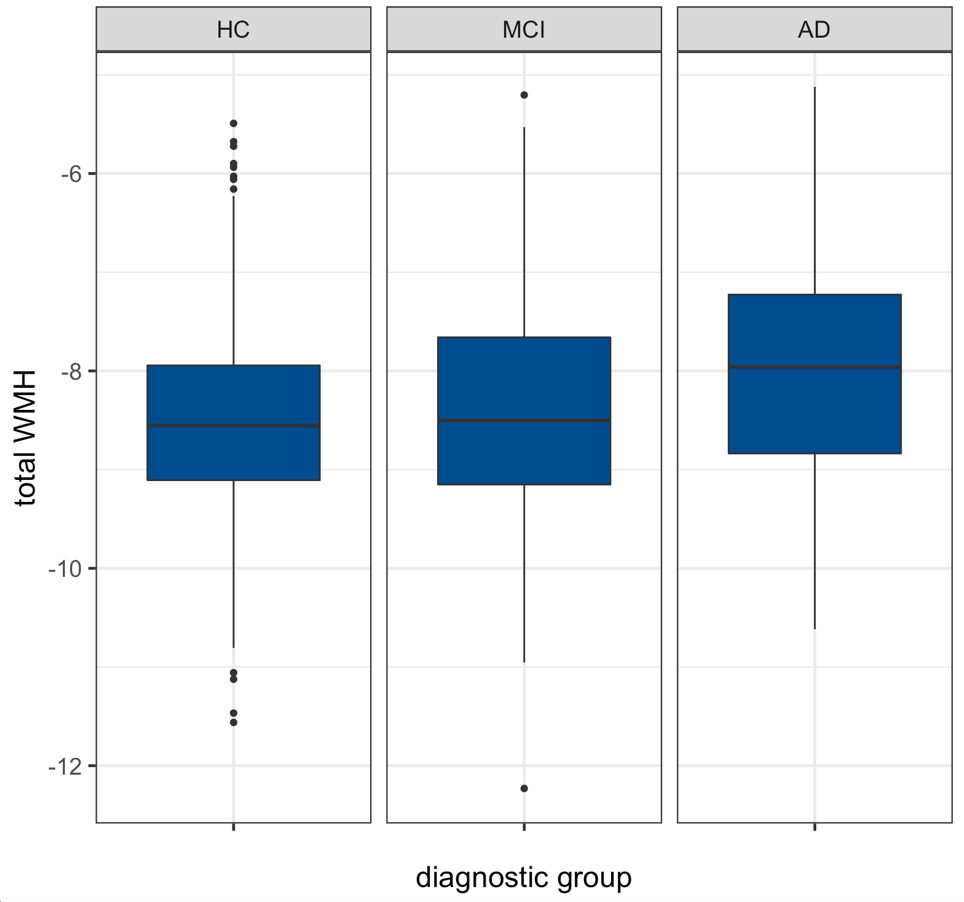

**Figure S1: Boxplots of natural-log transformed total WMH normalised by ICV for all subjects across diagnostic groups, healthy control (HC), mild cognitive impairment (MCI) and Alzheimer’s disease (AD)**

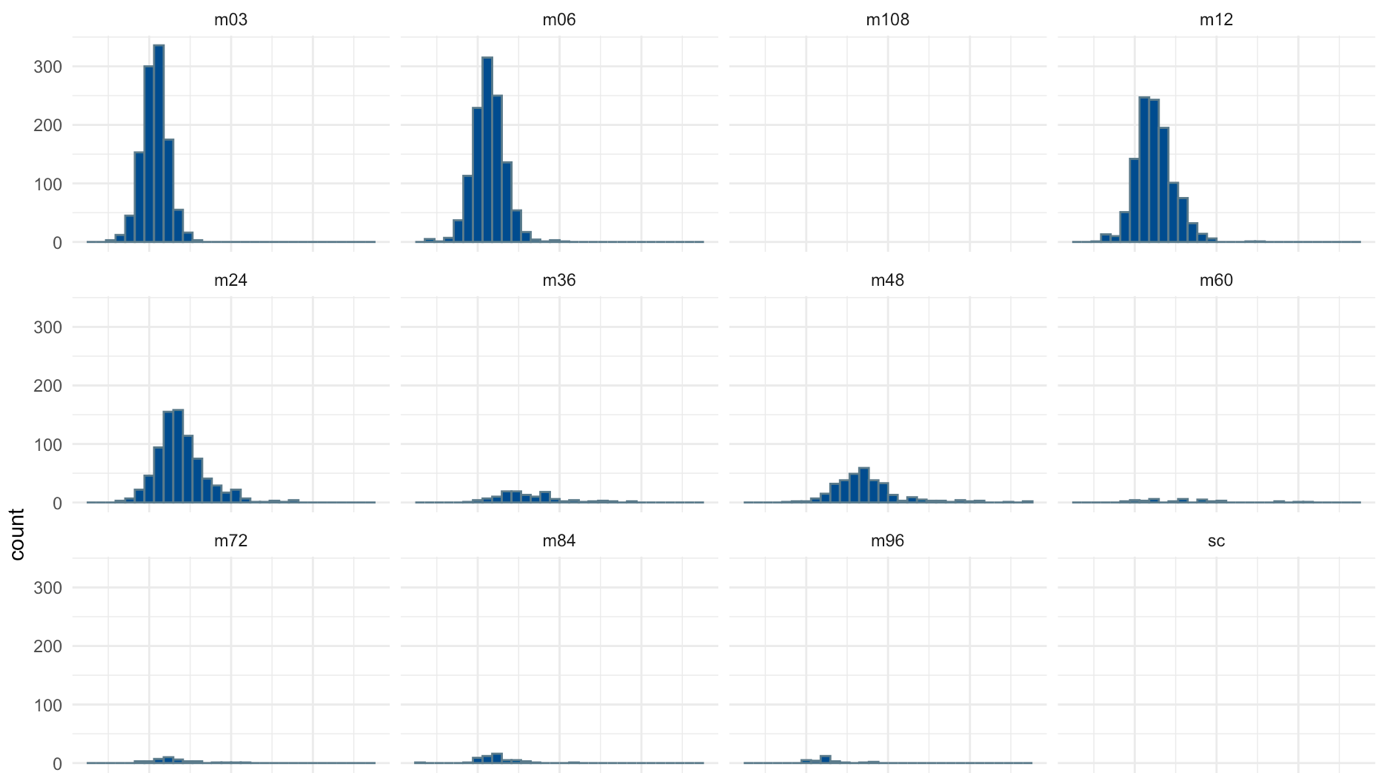

**Figure S2: Number of longitudinal BSI scans in CORE DATASET 1.**

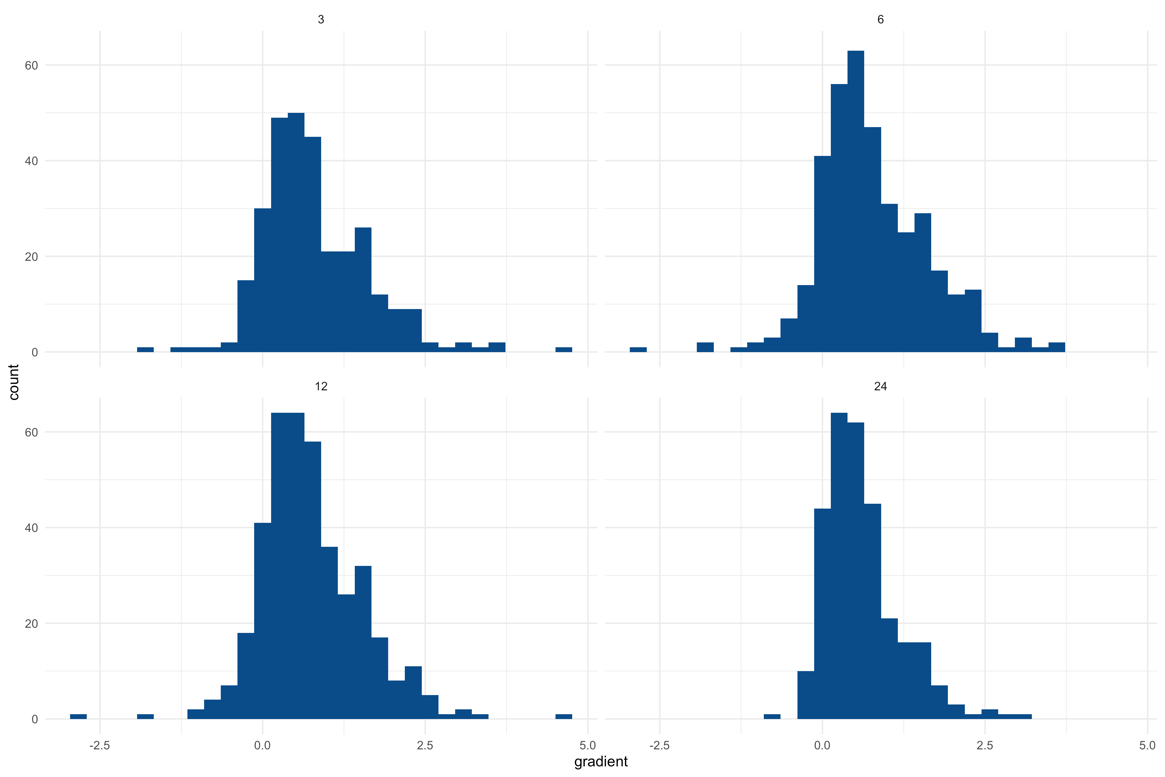

**Figure S3: Number of longitudinal BSI in CORE DATASET 2 over months 3, 6, 12 and 24.**

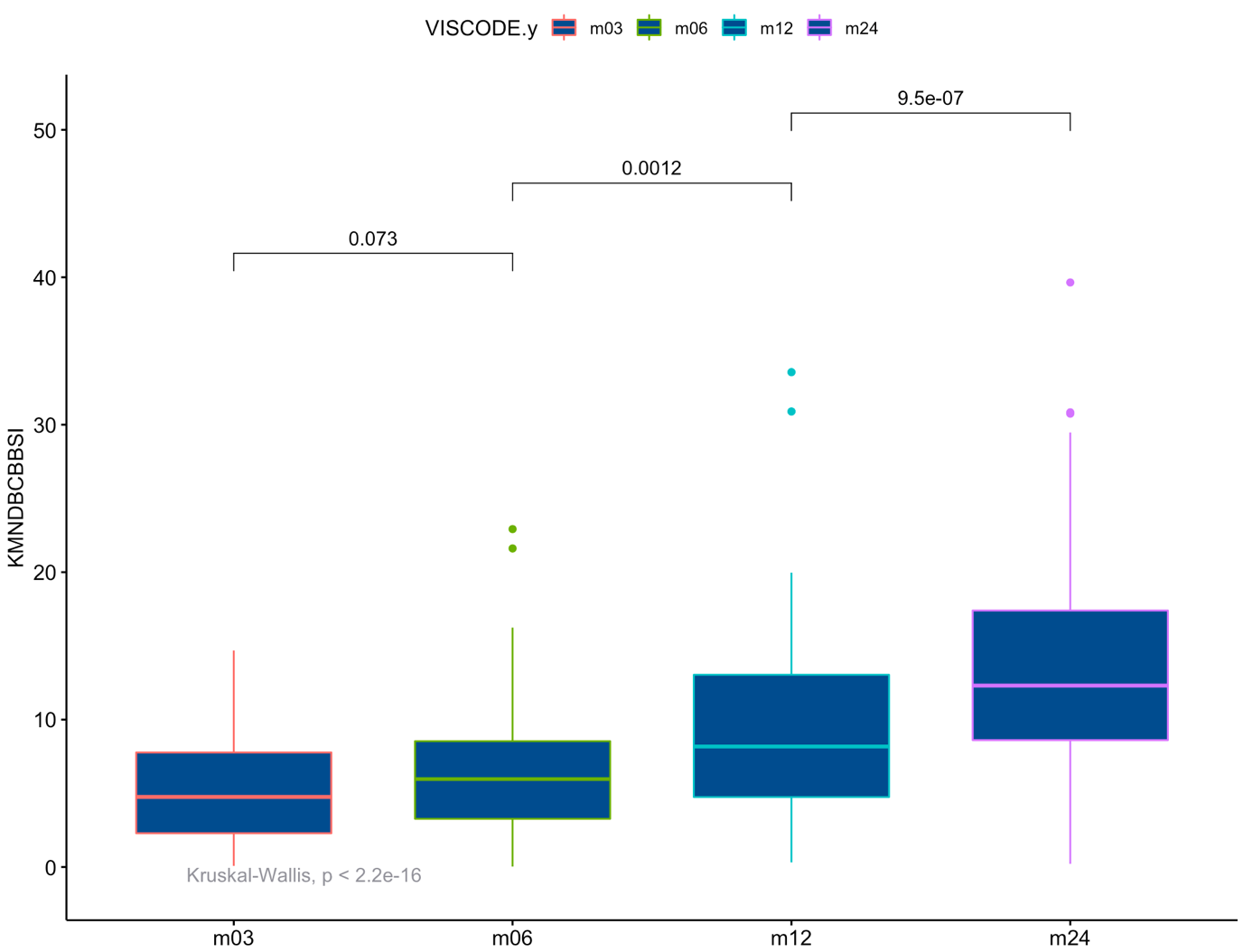

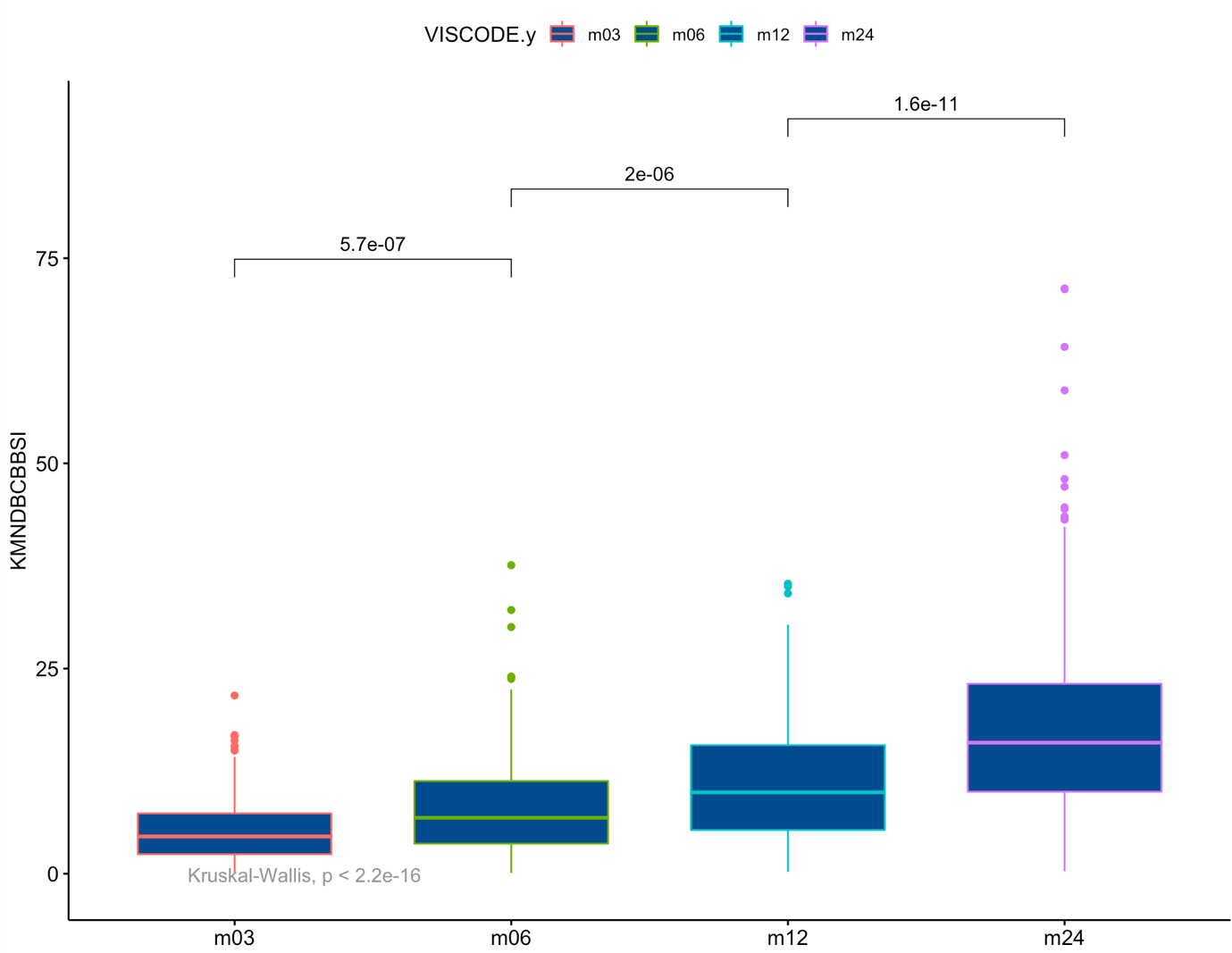

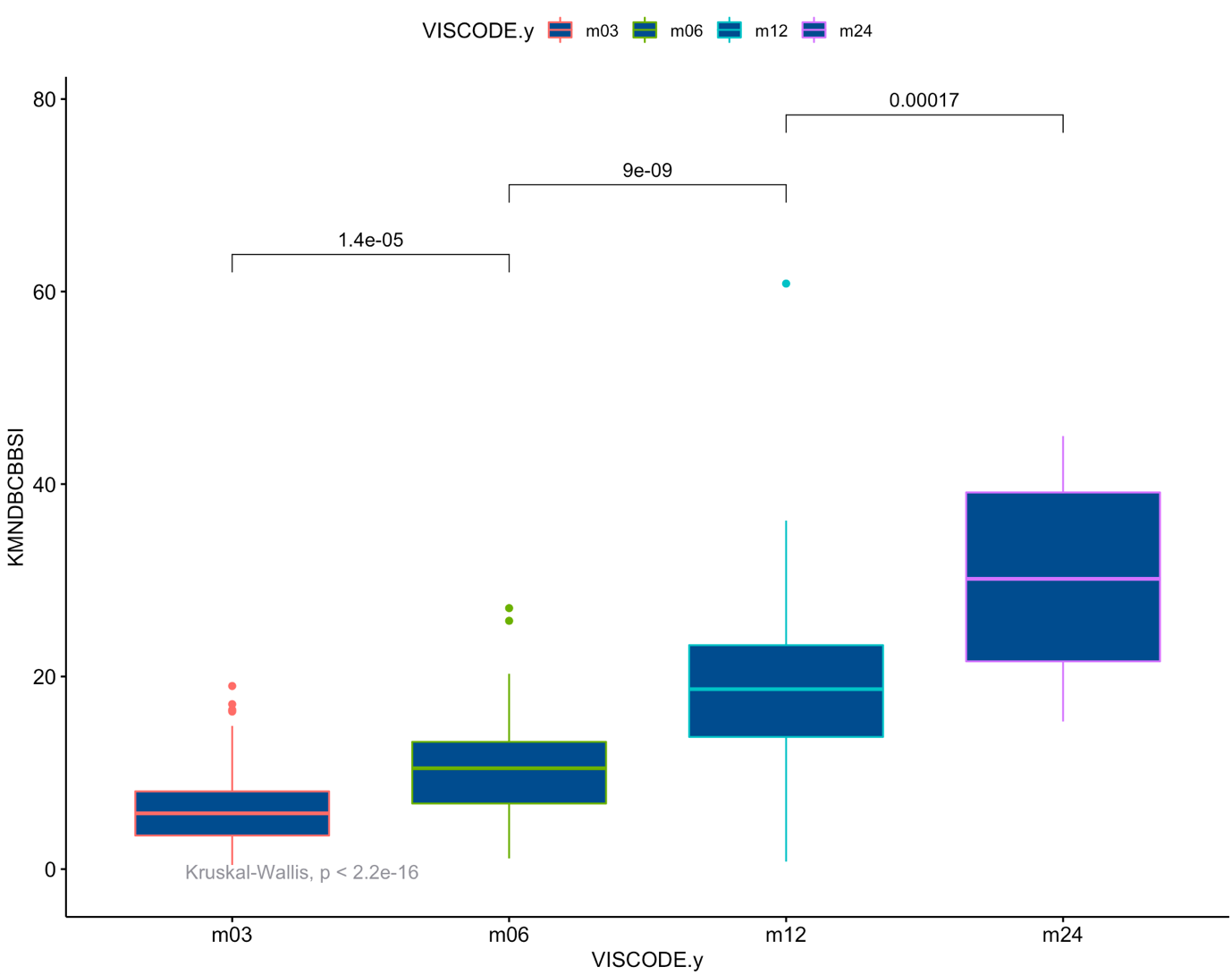

**Figure S4: Kruskal-Wallis multiple pairwise comparisons between scan months over diagnostic groups HC (top), MCI (middle) and AD (bottom).**

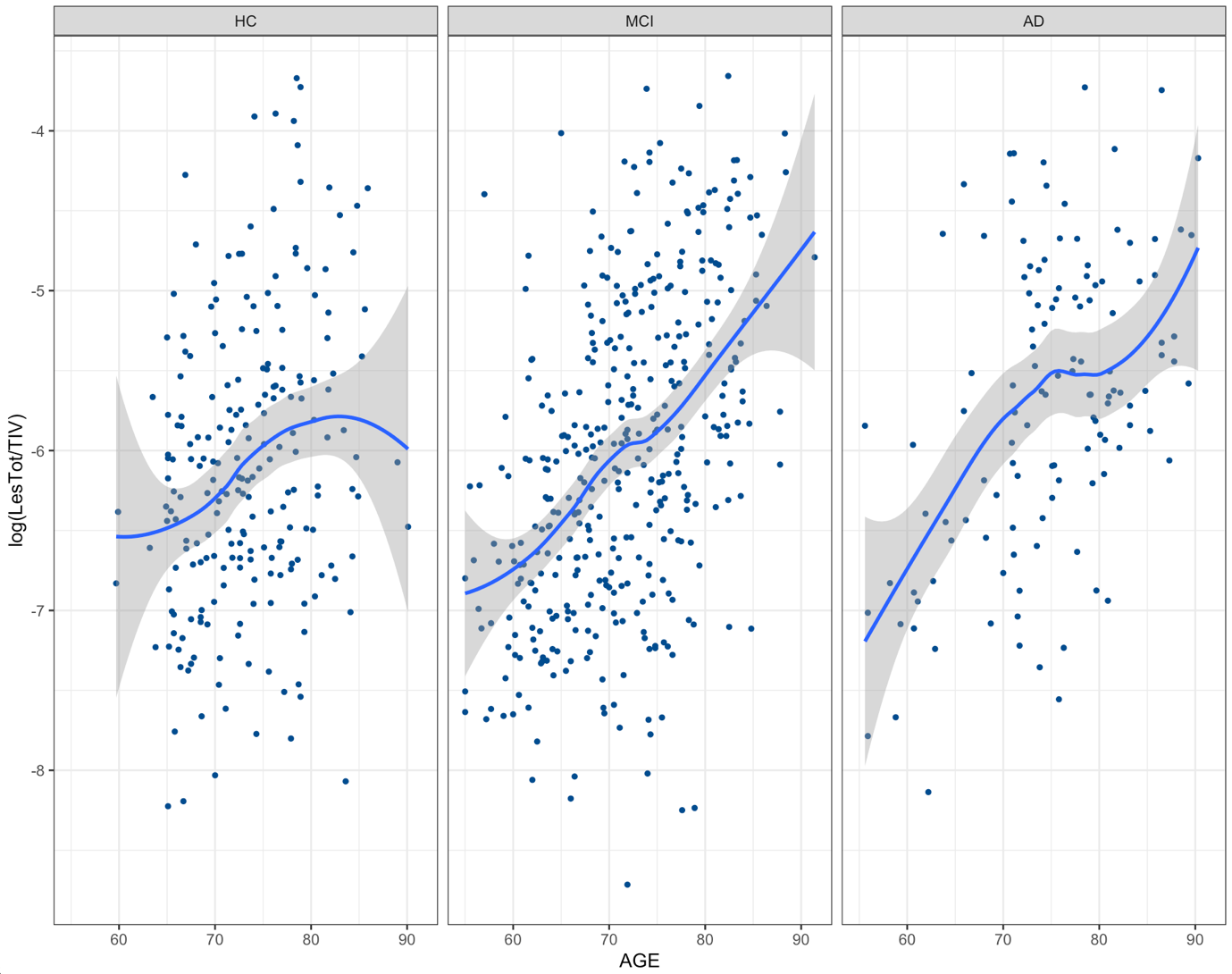

**Figure S5: Correlation between age and WMH load.** Natural log transformed normalised (for total intercranial volume) total WMH versus age (years) across diagnostic groups for CORE DATASET 1. Overlayed local polynomial regression fit with smoothing.

**Table S1: Numbers of scans per diagnostic group per time scan was take for CORE DATASET 2**

|  | HC | MCI | AD | total |
| --- | --- | --- | --- | --- |
| month 3 | 92 | 171 | 65 | 328 |
| month 6 | 93 | 241 | 67 | 401 |
| month 12 | 105 | 268 | 65 | 438 |
| month 24 | 132 | 187 | 15 | 334 |
| total | 422 | 867 | 212 | 1501 |

**Table S2: Tukey multiple comparisons of means for natural-log transformed total WMH normalised by ICV stratified by sex across diagnostic groups, healthy control (HC), mild cognitive impairment (MCI) and Alzheimer’s disease (AD)**

|  | female (adjusted p-value) | male (adjusted p-value) |
| --- | --- | --- |
| MCI-HC | 0.99 | 0.41 |
| AD-HC | 0.05 | 0.004 |
| AD-MCI | 0.03 | 0.04 |

**Table S3: Results from CORE DATASET 2.** T-values (p-values) for a range of KN-BSI whole brain atrophy over scans at months 3, 6, 12 and 24 with respect to natural log of regional WMH (at baseline or month 0) - frontal (F), parietal (P), occipital (O), temporal (T) and basal ganglia + thalami + infratentorial (BGIT); linear regression with confounders for age, sex, education, APOE4 burden and five principal components of population structure (including ICV confounder) across diagnostic groups.

|  | HC | |  | | | | | MCI | | |  |  | |  | |  | |  | AD | |  | |  | |  | |
| --- | --- | --- | --- | --- | --- | --- | --- | --- | --- | --- | --- | --- | --- | --- | --- | --- | --- | --- | --- | --- | --- | --- | --- | --- | --- | --- |
| KNBSI | F | P | | O | T | BGIT | F | | P | O | | | T | | BGIT | | F | | | P | | O | | T | | BGIT |
| month3 | 2.1 (0.04) | 1.8 (0.07) | | 2.2 (0.03) | 1.3 (0.2) | 1.3 (0.2) | 1.6 (0.1) | | 1.6 (0.1) | 1.5 (0.14) | | | 0.5 (0.65) | | 1.5 (0.14) | | 3.1 (0.002) | | | 2.3 (0.02) | | 3.1 (0.002) | | 1.2 (0.23) | | 2.2 (0.03) |
| month6 | 1.8 (0.07) | 2.1 (0.03) | | 1.8 (0.07) | 1.0 (0.3) | 2.1 (0.04) | 2.4 (0.02) | | 2.6 (0.01) | 2.3 (0.02) | | | 0.87 (0.38) | | 2.2 (0.03) | | 2.3 (0.02) | | | 2.6 (0.01) | | 2.4 (0.02) | | 0.59 (0.56) | | 2.4 (0.02) |
| month12 | 4.5  (9.4e-06) | 3.7 (0.0002) | | 3.8 (0.0001) | 2.9 (0.004) | 4.5 (8.3e-06) | 2.7 (0.008) | | 2.3 (0.02) | 2.6 (0.008) | | | 1.1 (0.27) | | 2.1 (0.04) | | 3.2 (0.002) | | | 2.6 (0.01) | | 2.9 (0.004) | | 2.0 (0.05) | | 2.9 (0.004) |
| month24 | 2.9 (0.003) | 2.8 (0.005) | | 2.6 (0.01) | 1.6 (0.2) | 2.8 (0.005) | 1.5 (0.1) | | 1.1 (0.27) | 1.6 (0.12) | | | 0.4 (0.7) | | 1.4 (0.16) | | 0.49 (0.63) | | | 0.03 (0.98) | | 0.2 (0.87) | | -0.45 (0.65) | | 0.78 (0.44) |
